# Frequency of Hepatitis B, C and HIV infections among transfusion-dependent Beta Thalassemia patients in Dhaka

**DOI:** 10.1101/2020.04.28.20079764

**Authors:** Golam Sarower Bhuyan, Aftab Uz Zaman Noor, Rosy Sultana, Farjana Akther Noor, Nusrat Sultana, Suprovath Kumar Sarker, Md Tarikul Islam, Md. Abu Sayeed, Md. Imam Ul Khabir, A K M Ekramul Hossain, Syeda Kashfi Qadri, Syed Saleheen Qadri, Firdausi Qadri, Kaiissar Mannoor

## Abstract

Transfusion transmitted infections (TTIs) have remained a major deterrent to public health, particularly among the patients with transfusion-dependent Beta thalassemia in developing countries. Although proper donor selection through adoption of WHO-advised infection panel has lowered the rate of infections, the multi-transfused patients are not free of risk. The present study screened 148 transfusion-dependent Beta thalassemia patients to determine the frequency of HCV, HBV and HIV using ELISA method. Among these patients, infected cases with HCV, HBV and HIV were 13.51%, 3.37% and 0%, respectively. Moreover, 2% of the patients had co-infections with both HBV and HCV. The percentage of infections in the patients with frequent transfusion interval (≤30 days) was significantly higher (P<0.0005) than that in the patients with less frequent transfusion intervals (>30 days). Immunochromatography (ICT)-based rapid test kits are usually used to screen and confirm these infections in the blood of the patients. However, ICT-based tests are not sensitive enough to detect the infections. So, a combination of both Nucleic Acid testing (NAT) and serological testing are suggested to significantly reduce the risk of viral infections during blood transfusion.

## Introduction

Annually, million units of blood are collected from the donors worldwide as the blood transfusion is integral to management of patients suffering from diverse diseases, particularly hematological disorders. From a record of 2013, it has been seen that there were more than 112 million units of blood donation all over the world that year.^1^ Accordingly, transfusion transmitted infections (TTIs) continue to be a major public health issue in many parts of the world and multi-transfused patients of Thalassemia (a group of inherited hemoglobinopathies caused by mutations in the beta globin chain of hemoglobin) are at a particularly increased risk of TTIs.^2,3^ These inherited blood disorders generally occur in the countries of thalassemia belt including Mediterranean and portions of West Africa, North Africa, Middle East and South Asian countries including Bangladesh, India, and Sri Lanka.^4^ As the patients with thalassemia, especially the patients with beta thalassemia major (BTM) and a group of patients with Hemoglobin E-beta thalassemia (EBT) are transfusion-dependent and these patients are very much prone to transfusion-transmitted viral infections. After heart failure, viral infections are the second most common cause of mortality and the foremost cause of morbidity among the patients with thalassemia followed by patients with bacterial and parasitic infections.^5^ Although hepatitis B virus (HBV), hepatitis C virus (HCV), human immunodeficiency virus (HIV), West Nile virus (WNV), human T cell lymphotropic viruses I, II (HTLV-I/II) had been reported most frequently, HCV and HBV had been known as the most prevalent etiological agents of chronic viral hepatitis and hepatocellular carcinoma among the thalassemia patients.^6^

In the developed countries, the TTIs risks have been virtually eliminated because of the careful and efficient screening approaches.^7^ However, challenges in developing countries span the entire blood safety chain from donor selection to post-transfusion surveillance and these risks are really a burning question for public health development.^8^ Bangladesh lies in the global thalassemia belt with around 9,176 new thalassemic patients each year^9^ and the patients with transfusion dependent BTM and EBT undergo regular transfusions with varying degrees of frequencies of transfusions in every 7 to 120 days. However, although it is mandatory, World Health Organization (WHO)– recommended standards for donor blood screening are not always followed in developing countries like Bangladesh due to resource limitations and occasional lack of consciousness.^10^ The patients with transfusion dependent BTM and EBT in Bangladesh not only suffer from various complications for the disease itself, but also due to the lack of proper screening methods of donor blood, and thus transfusion-transmitted infections are threatening as ‘Silent killers’.^11^

The minimization of the risks of TTIs highly depends on selection of safe blood donors through screening of the blood using sensitive methods for pathogen detection and such precaution reduces the risk of allogenic blood transfusion. However, transmission of diseases still occurs if there is the inability of the test method to detect the disease in the ‘pre-seroconversion’ or ‘window’ phase of their infections, immunologically variant viruses, non-seroconverting chronic or immune–silent carriers and laboratory testing errors.^12^ In the present study, the patients with thalassemia used to visit Bangladesh Thalassemia Samity and Hospital for routine blood transfusion. In the hospital, immunochromatographic tastings (ICTs) were usually performed to detect the presence of a number of common infections in the blood of donors before every transfusion. However, these ICT-based rapid diagnostic test (RDT) kits have not been validated using confirmatory approaches, such as ELISA or molecular methods and usually the ICT-based methods have repeatedly demonstrated low sensitivity and specificity to detect the major TTIs.^12,13^ The present study investigated the frequency of HBV, HCV and HIV infections among transfusion-dependent beta thalassemia patients of the Thalassemia Samity and Hospital using ELISA method with the aim to explore the efficacy of RDT based screening method.

## Materials and Methods

### Study participants

The study enrolled a total of 148 patients with transfusion-dependent Beta thalassemia over a period of 7 months from August 2017 to February 2018. These patients were in the age range of 2-46 years (90 patients were <18 years). They used to visit Bangladesh Thalassemia Samity and Hospital for follow up examinations and blood transfusions. Both EBT and BTM patients participated in the study and they were all differentially diagnosed by Hemoglobin capillary electrophoresis. The study also included 30 healthy controls (aged 2-46 years) who did not have any previous history of hemoglobinopathies or related blood disorders. Prior to sample collection, a written informed consent was obtained from the adults as well as from the legal guardians of the children (both for the patients and the controls). About 5.0 milliliters of blood were collected from each participant in serum tubes using the standard venipuncture method and serum was separated from the collected blood. In case of patients, blood was collected before transfusion. The study was approved by National Ethics Review Committee (NERC) of Bangladesh Medical Research Council (BMRC), Dhaka, Bangladesh.

### Donor blood screening

The Bangladesh Thalassemia Samity and Hospital routinely use ICT kits (EXcEL®, USA) for detection of HBV, HCV and HIV in the blood of donors before transfusion. The donor blood was usually arranged from the hospital services or by the guardian of the patients through their personal contacts or from various blood banks. Every ICT-dependent suspected donor was usually excluded from the transfusion process.

### Detection of HBV, HCV and HIV in the blood of patients

As the presence of Hepatitis B surface antigen (HBsAg) in the serum is a good indicator of HBV infections, Bioelisa HBsAg 3.0 kits (Biokit, Barcelona, Spain) were used to detect HBsAg. Bioelisa HCV 4.0 (Biokit, Barcelona, Spain) was used for qualitative detection of anti-HCV antibodies in the sera of the patients. The HIV 1/2/O Antigen/Antibody EIA Test Kit (Acon Laboratories, CA, USA) was used for the qualitative enzyme immunoassay-based detection of HIV-1 P24 antigen and total antibodies (IgG, IgM and IgA) to HIV-1, HIV-2, and/or Subtype O in human serum or plasma. These tests were performed in accordance with the manufacturers’ instructions provided with the kits. The color intensity of the final reaction mixture was measured with an ELISA reader (LabSystems Multiskan, USA) at 450 nm.

### Statistical analysis

We performed Two-tailed chi-square test (two by two table) using http://www.openepi.com/ to calculate the frequency of infections among the two transfusion interval groups (≤30 days and >30 days). Estimation of the relative risk ratio (RR) and odds ratio (OR) were done at 95% confidence interval (CI). A P-value of less than 0.05 was considered significant.

## Results

### Demographic characteristics of the patients

In this study, 53.38% of the participants were males and 46.62% were females. There were both BTM and EBT patients among the participants. We recorded different demographic information of the patients like name, address, gender, height, weight, BMI as well as transfusion-related information like age of first transfusion, transfusion interval and number, splenectomy etc. The baseline information of the patients is given in the Table 1.

**Table 1.** Baseline information of the patients enrolled in this study

| Parameters |  |
| --- | --- |
| Gender | Male, N=79 (53.38%); Female, N=69 (46.62%) |
| Thalassemia type | EBT, N=90 (60.81%); BTM, N= 58 (39.18%) |
| Age (years), mean ± SD | 17.15 ± 9.33 |
| BMI (kg/m<sup>2</sup>), mean ± SD | 17.39 ± 3.32 |
| Transfusion interval (days), mean ± SD | 33.68 ± 29.98 |
| Transfusion number, mean ± SD | 228.32 ± 189.85 |
| Splenomegaly (cm), mean ± SD | 6.94 ± 3.09 |
| Splenectomy | N=21 (14.18%) |

2 (Abbreviations: EBT= HbE Beta Thalassemia, BTM= Beta Thalassemia Major, SD= Standard Deviation, BMI= Body Mass Index)

### Trend of infections among the patients

A total of 22 patients (14.86%) with thalassemia were found to be infected (Male: N=12, 54.54% and female: 10, 45.45%). Among these 22 infected thalassemia patients, we identified 19 (12.83%) patients infected with either HBV or HCV and 3 (2%) of them had coinfections with both HBV and HCV. Also, the number of patients infected with HCV (N=20, 13.51%) was higher than that of HBV (N=5, 3.37%) (Table 2). Moreover, none of the patients was infected with HIV and all the healthy controls were negative for HBV, HCV and HIV (data not shown).

**Table 2.** Frequency of viral mono-infection and co-infection among the transfusion-dependent EBT and BTM patients

| Thalassemia<br>type | Infection Type |  |  |  | No. of<br>Total<br>infected<br>(%) |
| --- | --- | --- | --- | --- | --- |
|  | No. of patients<br>with HBV and<br>HCV (%) | No. of<br>patients with<br>HBV (%) | No. of<br>patients with<br>HCV (%) | No. of<br>patients with<br>HIV (%) |  |
| EBT (N=90) | 1(1.11) | 2(2.22) | 11(12.22) | 0(0) | 14(15.55) |
| BTM (N=58) | 2(3.44) | 0(0) | 6(10.34) | 0(0) | 8(13.79) |
| Total (N=148) | 3(2.02) | 2(1.35) | 17(11.48) | 0(0) | 22(14.86) |

- 1 (Abbreviations: EBT= E Beta Thalassemia, BTM= Beta Thalassemia Major, HBV= Hepatitis B Virus, HCV= Hepatitis C Virus, HIV= Human - 2 Immunodeficiency Virus)

### Relation of infection frequency with transfusion interval

We categorized the patients into two groups based on their transfusion interval. One of these groups had the transfusion interval ≤30 days, whereas the other had the transfusion interval of >30 days. In the more frequent (≤30 days) transfusion group, the number of virally infected patients (N = 18, 25.35%) was found to be significantly higher (P-value < 0.0005, RR = 1.945 [1.464 −2.585], OR = 6.198 [1.983 −19.37]) than that of the less frequent (>30 days) transfusion group (N = 4, 5.19%) (Figure 1).

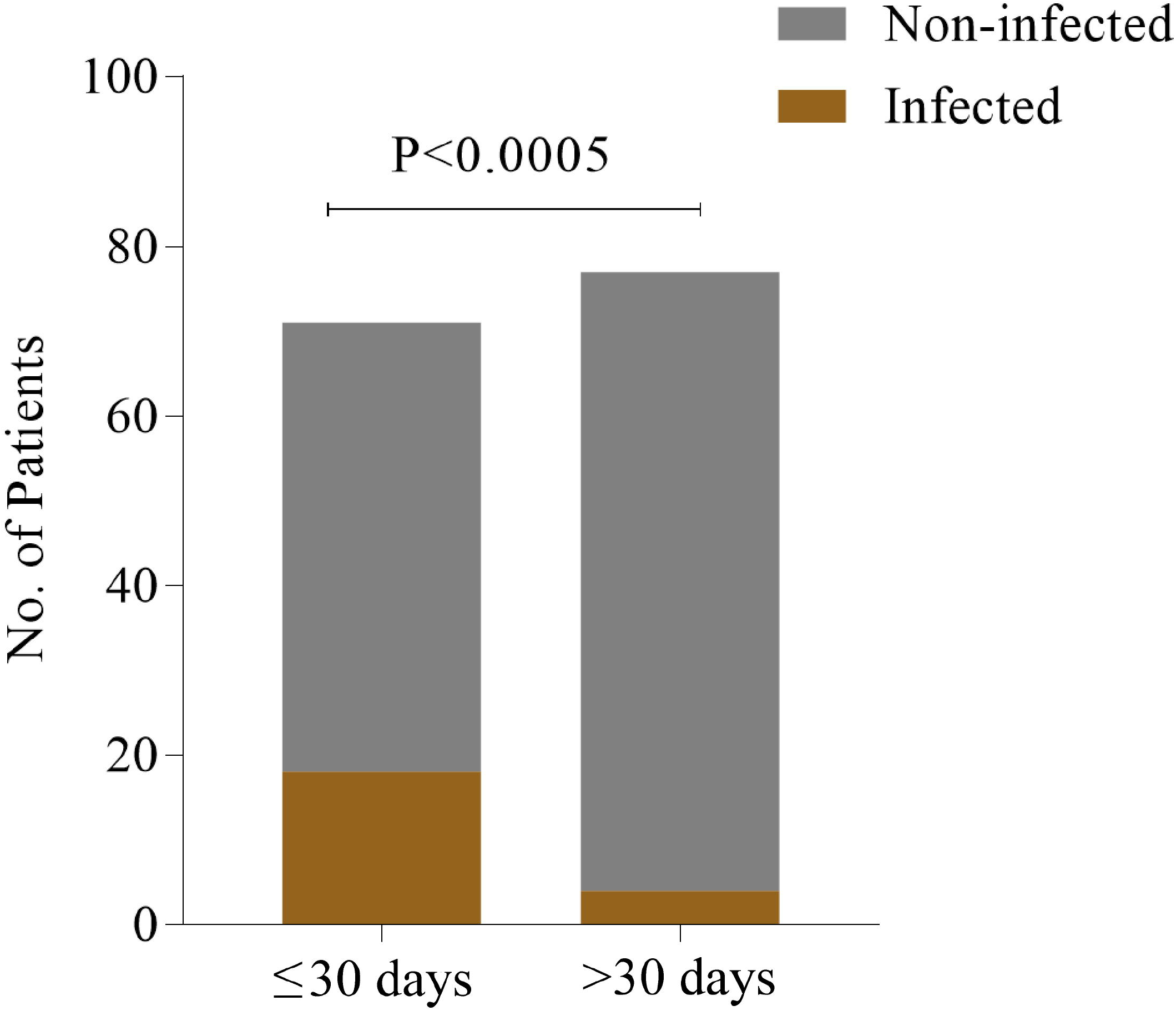

## Discussion

In Bangladesh Thalassemia Samity and Hospital, effective screening processes for infections are done before blood transfusion for the proper selection of donors. However, still we could identify patients with either HBV or HCV or both. The most important question was why some of the patients got infected albeit standard screening processes were followed before every transfusion.

In case of HBV, there were some asymptomatic donors who were in the ‘window period’ (i.e. the early infection period when an immunologic test is non-reactive) without any expression of HBsAg. Also, people with occult HBV (absence of HBsAg and the presence of HBV DNA in liver tissues with or without HBV DNA in the serum) may remain undetected by traditional ICT kits.^14^ Previous study also showed that the sensitivity of ICT-based rapid tests was not higher enough to detect hepatitis status of a donor and these kits might not detect certain prevalent serotypes of HBV in any particular region. ^8^ Detection of the HBV DNA by implementation of nucleic acid testing (NAT) assays had been reported to shorten the core window period to reduce the residual risk. ^15^. In this study, the number of patients infected with HCV was higher than that of HBV. It should be noted here that although there were HBV vaccine coverage to some extent in Bangladesh, there were still no HCV vaccine available yet.^16^ This may explain why there were more thalassemia patients infected with HCV than with HBV. Rate of infection was found to be significantly higher in the patients who had to undergo frequent transfusions which indicates that multi-transfused thalassemia patients are more prone to infections. This is compatible with previous findings that demonstrated that multiple transfusions receiving for a long period of time might cause immunomodulation resulting in susceptibility to infections.^7^

In Bangladesh Thalassemia Samity and Hospital, sometimes ELISA methods are applied as a confirmatory test after the donors are suspected as positives by ICT approach. Nevertheless, previous study had shown that ELISA only could detect HBsAg, whereas real-time PCR could represent infection status by detecting HBV-DNA. ^8^ On the other hand, recombinant immunoblot assay (RIBA) had more accuracy in detecting HCV.^16^ The multi-transfused patients with thalassemia may face increased immune dysfunction in the presence of iron overload following splenectomy, which makes them more susceptible to infections.^17^ Moreover, in case of emergency, there is a common practice in Bangladesh of risky blood donations by professional donors without any kind of testings who are mostly drug abusers.^18^ Although nowadays an effective awareness in safe blood transfusions and iron chelation therapy have made the morbidity rate of patients with transfusion-dependent Beta thalassemia lower, new complications like hepatocellular carcinoma are taking hold in the patients with thalassemia, which may be due to carcinogenicity of iron overload and chronic infections.^19^ Specially, the thalassemia patients who become co-infected with both HBV and HCV are at a greater risk of cirrhosis and hepatocellular carcinoma compared to the mono-infected patients.^20^

Hence, there is no alternative of making blood transfusion process safer by proper selection of voluntary healthy blood donors, along with NAT which reveals viral agents earlier in the ‘window period’ than other immunoassays. Additionally, screening of HBV-DNA and HCV-RNA needs to be practiced regularly to avoid any kind of risks during blood transfusions.^17^ Studies in India, China and Saudi Arabia had reported that NAT-based detection methods for HCV and HBV could tremendously improve the efficacy of screening for protecting blood recipient from TTIs.^21-23^ Furthermore, introduction of HBV NAT in the USA, along with the HBV vaccination policy made a substantial contribution to blood transfusion safety and decreased residual risk of HBV infections.^24^ In the UK, NAT has reduced the risk of HCV by 95% and that of HIV by 10%.^25^ However, in case of very low levels of viremia, NAT might not be effective enough to detect the infections. Despite the limitations, to ensure safe blood transfusion, combination of both NAT and serological testing may significantly reduce the risk of viral infections during transfusion and thus may improve the quality of life of the thalassemia patients.^6^

In this study, we could not compare between serological and NAT-based methods due to budget limitations. Such a comparison could have been a concrete evidence for urging NAT-based screening for blood transfusion in Bangladesh. Moreover, inclusion of other pathogenic organisms in the screening panel could have provided us more information on the severity of the current situation.

## Conclusion

The patients requiring frequent blood transfusion are more prone to HCV and HBV infections, which can be attributed to unsafe transfusions. To give these patients good quality of life from these preventable complications, efficient screening method for proper donor selection should be adopted in Bangladesh. In addition, awareness should be built to ascertain a safe transfusion. Such practices will ensure a safe blood transfusion for the patients suffering from other diseases and save them from unwanted life-threatening complications.

## Data Availability

All relevant data are within the paper. Further information is available from the authors on request.

## Acknowledgements

We thank the clinicians and staff of Bangladesh Thalassemia Samity and Hospital for their assistance in specimen collection.

